# A computationally efficient method for dimensionality reduction in multi-channel spectral CT

**DOI:** 10.1101/2024.02.18.24302905

**Authors:** Olivia F. Sandvold, Roland Proksa, Heiner Daerr, Amy E. Perkins, Kevin M. Brown, Thomas Koehler, Ravindra M. Manjeshwar, Peter B. Noël

**Affiliations:** Department of Radiology, Perelman School of Medicine, University of Pennsylvania, Philadelphia, USA; Department of Bioengineering, University of Pennsylvania, Philadelphia, USA; Philips Innovative Technologies, Hamburg, Germany; Philips Healthcare, Cleveland, USA

**Keywords:** computed tomography (CT), quantitative imaging, spectral CT, DECT, kVp-switching, iodine contrast, hybrid spectral CT, multi-energy CT

## Abstract

**Objective:** Multi-channel spectral CT technology holds the promise of measuring incoming X-rays across various energy spectrums, significantly enhancing iodine imaging performance. Nonetheless, it necessitates the reduction of data dimensions, such as condensing multiple energy bins, to ensure processing times remain within clinically acceptable limits and to maintain compatibility with traditional two-dimensional material decomposition methodologies. This study presents an optimization strategy designed to efficiently integrate multi-channel spectral CT data, aiming to minimize the anticipated noise in the iodine domain across different system parameters influencing channel content.

**Approach:** The Cramer-Rao lower bound of variance (CRLB) was used to estimate the iodine domain noise for four two-input weighting schemes including kVp-switching compared to a four-input material decomposition noise estimate. Two of the four weighting schemes were optimized to contain weights that minimized the expected CRLB noise. A model of rapid kVp-switching x-ray tube and dual-layer detector were used to simulate acquisitions of a hybrid, combined technology multi-channel spectral CT system. The impact of the duty cycle ratio, or proportion of high kVp to low kVp, and the phantom size ranging from adult to pediatric were investigated.

**Main Results:** The two-input weighting schemes with varying amounts of each spectral channel, optimized for low iodine noise, showed consistent low estimated noise performance within 0.27% difference to the ideal, four-input material decomposition results for all tested duty cycles in a standard adult-sized 300 mm water phantom. These schemes were always superior to kVp-switching regardless of duty cycle or phantom size. In the pediatric (150 mm) and large adult (400 mm) phantom cases, the two-input weighted schemes were within 1% difference to the ideal four-input noise estimator results on average, across all tested duty cycles.

**Significance:** Advanced dimensionality reduction techniques, such as those presented in this work, that merge multi-channel spectral CT data illustrate a potential in improvement the accuracy of iodine contrast material measurement, while filling clinical specifications. This study also indicates the significant role that hybird spectral CT technologies could play in elevating the reliability of quantitative CT.

## INTRODUCTION

Spectral computed tomography (CT) enables quantitative imaging results providing physiologic and functional insights to enhance CT diagnostic utility^1,2^. The additional spectral results including monoenergetic images and material-specific maps allow for increased lesion detection, decreased noise, and tissue analysis^3–6^. Spectral CT acquisitions provide the greatest clinical benefit with high accuracy in separation and quantification of material composition and radiation dose exposures set to be equivalent or lower than conventional CT imaging. From a technological standpoint, there are several approaches to enable clinical spectral CT^7,8^. These include: dual-source geometry^9^, slow/spin-spin and rapid kVp-switching (kVp-s)^10,11^, spectral detectors^12,13^ (either energy-integrating or photon-counting), and twin-beam filters^14^. There exist some physical constraints for each of these implementations, particularly relating to spectral separation^15^, susceptibility to motion artifacts^16^, electronic noise and scatter^17^, and spectrum modeling mismatch^18^, all which generate noise and produce bias. In ultra-low dose scenarios, increased noise may further increase bias in spectral results^19^. This is unacceptable for quantitative tasks such as measuring iodine uptake in oncological lesions where high sensitivity of iodine estimation is required for evaluation of therapy response and correct identification of metastases is dependent on accurate CT characterization^6,20^.

It has been demonstrated in prior work that CT systems designed to produce more than two spectral datasets can yield lower bias and lower noise, particularly for low concentration iodine^21–24^. One way to obtain additional spectral information is through use of a photon-counting detector (PCD) with three or more energy threshold bins^25^. Another possibility is to use a hybrid spectral CT system designed to address hardware limitations by combining dual-energy technologies. The ability to select a subset of the acquired photon counts is a desirable trait of both hybrid and PCD technology. The current implemented reconstruction process on the commercially available PCD equipped CT scanner produces three energy bins where the lowest energy bin is reserved for removal of electronic noise. However, several publications have shown that generation and use of an increased number of energy segregated data channels improves spectral performance and enables three-material decomposition^26–28^. Design optimization of such systems has yet to be further explored and implemented into a clinical workflow. There remain pertinent questions regarding how to best integrate multi-channel data into material decomposition processes under clinical processing time and resource limitations. Obstacles to utilization of multiple channel information for spectral imaging results include costly slip-ring upgrades, processing hardware, raw data storage, and increased computational reconstruction complexity. Further, the performance of these systems should significantly improve upon existing solutions.

In this work, we introduce practical weighting schemes to minimize complexity and expected noise in material decomposition from projections acquired through multi-channel CT scans. In the absence of a K-edge material, the overwhelming majority of clinical scenarios only require a two-material decomposition while additional information from more than two input channels has potential to enhance spectral performance in terms of noise and bias reduction. Therefore, our computationally efficient weighting schemes permit a reduction in projection space dimension, enabling the utilization of established decomposition techniques at ideal noise performance. Specifically, we utilize spectral data simulated from a rapid kVp-switching and dual-layer hybrid CT system to demonstrate the weighting schemes for optimal iodine quantification noise. By reducing projection space data channels from four to two, we benefit from decreased hardware requirement upgrades, capability to use existing clinical processing workflows, and most importantly, fine-tuned selection of spectral data from multiple energy domains.

Material decomposition approaches in spectral CT determine the mass attenuation contributions from either the photoelectric effect and Compton scatter interactions or between two clinically relevant material basis functions, e.g., water and iodine, to estimate underlying tissue properties^1,29^. The Cramer-Rao lower bound (CRLB) to estimate basis image noise from single line-projection acquisitions was demonstrated first by Alvarez and Macovski and subsequently by others to optimize design parameters and estimate quantitative performance of unique CT system configurations^29–31^. One advantage of this statistical approach is that the CRLB can be extended to include more than two input spectra and two output basis material estimations. However, in a clinical CT system, increasing the number of spectral inputs or desired material decompositions is complex and increases computational burden. Therefore, we realistically constrain our material decomposition schemes to use only two spectral inputs composed from weighted sums of the multiple data channels acquired and to estimate water and iodine noise in a simplified phantom. The optimal contribution of each data channel is determined by minimization of the expected CRLB noise.

The foundation of this proposed weighting methodology builds upon the knowledge that excellent spectral separation is essential for high accuracy material quantification in spectral CT^1^. In photon-counting technology, spectral separation is a product of photon energy thresholding to create multiple energy bins. In comparison, the hybrid CT configuration considered here includes a dual-layer detector and kVp-s tube would produce four distinct “data channels” of energy-integrated signals provided by the combinations of high and low tube voltages and upper and lower detector layers. The spectral separation would arise from the discrimination of polychromatic spectra from two distinct tube voltages by the dual-layer detector. To both feasibly handle multi-channel data and optimize the addition of the data to a general spectral forward model, we introduce a weighting method to do the following: (1) implement a linear weighting scheme to combine the four channels into two new spectral inputs; (2) use the CRLB as an optimization function to identify optimal weights over varying scan parameters. We use a simulation to compare the noise estimate from our simplified 2-channel weighting schemes to an idealized 4-channel performance. In this paper, we show the potential for a multi-channel hybrid spectral CT system which outperforms the single technology two-energy systems and uses compressed spectral data to achieve on par noise statistics to the full-spectrum four-channel simulated model.

## METHODS AND MATERIALS

### Phantom materials

In this experiment, a simulated phantom was modeled by defining path lengths through water [1 g/mL] and iodine [10 mg/mL]. For an ‘average’ adult-sized patient, the path length was set to 300 mm of water to model a diameter of 300 mm. Our large adult patient was modeled with a 400 mm water path length while the pediatric case used a path length of 150 mm. This pediatric diameter roughly approximated the 50^th^ percentile of 2-year-old male and female children in the United States^32^. All iodine path lengths were set to 25 mm to represent a small lesion or vessel. For perfusion imaging, 10 mg/mL represents a moderate amount of iodine contrast that should be detectable by CT. Mass attenuation values were provided by the National Institute of Standards and Technology^33^.

### Scan parameters

The rapid kVp-switching tube emission output were modeled from a commercial tube (Philips Healthcare) operating in rapid switching mode between 140 and 80 kVp using manufacturer specified spectra estimating the number of photons/keV/mA/s/sr defined from 10 to 150 keV in 1 keV intervals^34^. Attenuation from tube housing, aluminum filtration, and a commercial bowtie filter was applied. The tube current was set based on patient diameter. For the 300 mm, or average adult-sized phantom setting, a current of 200 mA was used for the 140 kVp projection and 146 mA was used for the 80 kVp projection. The rotation time was set to 2 seconds with 2000 projections per tube voltage or 0.5 ms per exposure. The current difference accounts for the manufacturer’s expected decrease in emission current for ultrafast switching to lower tube voltage. Although the emitter temperature of the X-ray tube will stay constant, the field related emission will drop. **Table 1** shows the tube current used for each tube voltage for the pediatric and large adult phantoms.

**Table 1:** Simulation parameters. Simulation scan parameters to define each tube spectra, φ_*kVp*_(*E*), for the tested phantom sizes and duty cycle ratios. Each water phantom was simulated with an additional 25 mm of 10 mg/mL iodine.

| Simulation Parameters | High kVp | Low kVp |
| --- | --- | --- |
| Tube voltage [kVp] | 140 | 80 |
| Tube current [mA] |  |  |
| 150 mm phantom | 100 | 73 |
| 300 mm phantom | 200 | 146 |
| 400 mm phantom | 300 | 219 |
| Cycle time % | 10 | 90 |
|  | 25 | 75 |
|  | 50 | 50 |
|  | 75 | 25 |
|  | 90 | 10 |

The central ray of the tube spectra for each tube voltage, represented as φ_*kVp*_(*E*) in (1), were each multiplied by a general detector layer response, *D_layer_*(*E*), for the upper and lower scintillators to produce four corresponding data channels, *S*_*kVp*,*layer*_(*E*). The dual-layer detector responses contain the spectral sensitivities of the two detector layers with a central positioned geometry and were supplied by the manufacturer^34^. Note that the upper layer signal was mainly composed of low energy photon signal while the lower layer signal was primarily composed of response to high energy photons. The duty cycle, or ratio of percent of cycle time spent using 140 kVp tube voltage compared to 80 kVp in one pair of projections, was initially set to 50/50. In this experiment, one pair of 140 and 80 kVp projections were integrated over a period of 0.5 ms each. This yielded a total cycle time of 1 ms.

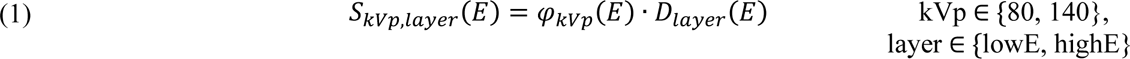

An estimated patient radiation dose per sr was given by 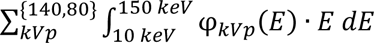, or the sum of estimated photon energy in one pair of high and low kVp incident spectra. We refer to this value as d_140+80_ while d_140_ and d_80_ represent the estimated sum of photon energy for the 140 and 80 kVp components, respectively. d_140+80_ was proportional to the airkerma for the scan. We remodeled the spectra φ_*kVp*_(*E*) to use the duty cycles 10/90, 25/75, 75/25, and 90/10 as generating the tube spectrum relied on the cycle time input. We then calculated the new estimated patient radiation dose and linearly scaled the new spectra to contain equivalent total photon energy sum (d_140+80_) to the initial 50/50 duty cycle estimated patient radiation dose. Therefore, we created a single pre-patient “dose” level across duty cycles. The 10/90 case represented favorable conditions where the 140 kVp component is much shorter in duration compared to the duration of 80 kVp tube voltage. The longer duration for the lower voltage provided an improved flux and noise balance between the two spectral channels that otherwise suffers from the strong flux decrease for lower tube voltages. This configuration may reduce absorbed radiation dose for certain patient habitus profiles. The 90/10 duty cycle represented a theoretical scan protocol necessary to image a large patient with higher demands on the total flux that can be generated with the high voltage.

### Weight scheme design

We used a two-input CRLB to model the two-channel material decomposition approach. To do this, we defined two new spectra using the available four data channels by selecting weights for each measurement channel indicating the amount of contribution from that spectra that should be included in the final decomposition for each input *i* (2). Summing over the linearly applied weighted channel spectra resulted in two new energy spectra, Φ_*i*_(*E*) (3). It then followed that the Beers-Lambert law was applied to obtain the expected total energy-integrating measurement for the new channels through pre-defined phantom materials *M* with linear attenuation coefficients *μ* and path lengths *X*, neglecting electronic noise.

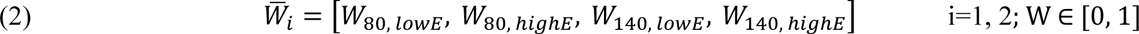

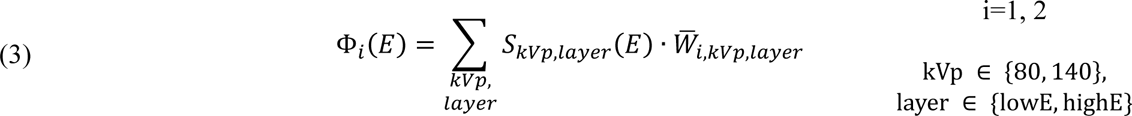

Supposing that there existed two potential states (zero or non-zero) for a single weight value, of which there were four for each weight matrix, and there were two weight matrices total, a total of 2^8^ possible combinations could have been implemented with this method. However, this number neglects removal of zero weight matrices which would be impractical to use. Realistically, the number of reasonable reduced dimension weight schemes remained within single digits given the spectral identities of each of the four channels. In this paper, two general two-input channel schemes were proposed (Figure 2) to simplify the number of possible optimization points. In the first scheme, the contribution of the low energy photons in the high kVp projection (denoted by *b*) combined with the contribution of a mix of 80 kVp dual-layer detector signals to form one new low energy input. The high energy photons (lower layer) signals in the high kVp projection served as the second CRLB input. Contrast this with the second scheme where the low energy photons in the high kVp projection (*c*) were added to the high kVp, high energy photons as the second CRLB input; the first CRLB input consisted of only low kVp signals. The *a* weight for both schemes was adjusted to contribute high energy photons from the 80 kVp projection to the first input to the CRLB. We excluded other combinations as they showed very poor noise performance. Using these weight notations, kVp-switching was modeled by setting *a=1, b=0,* and *c=1*.

**Figure 2:**
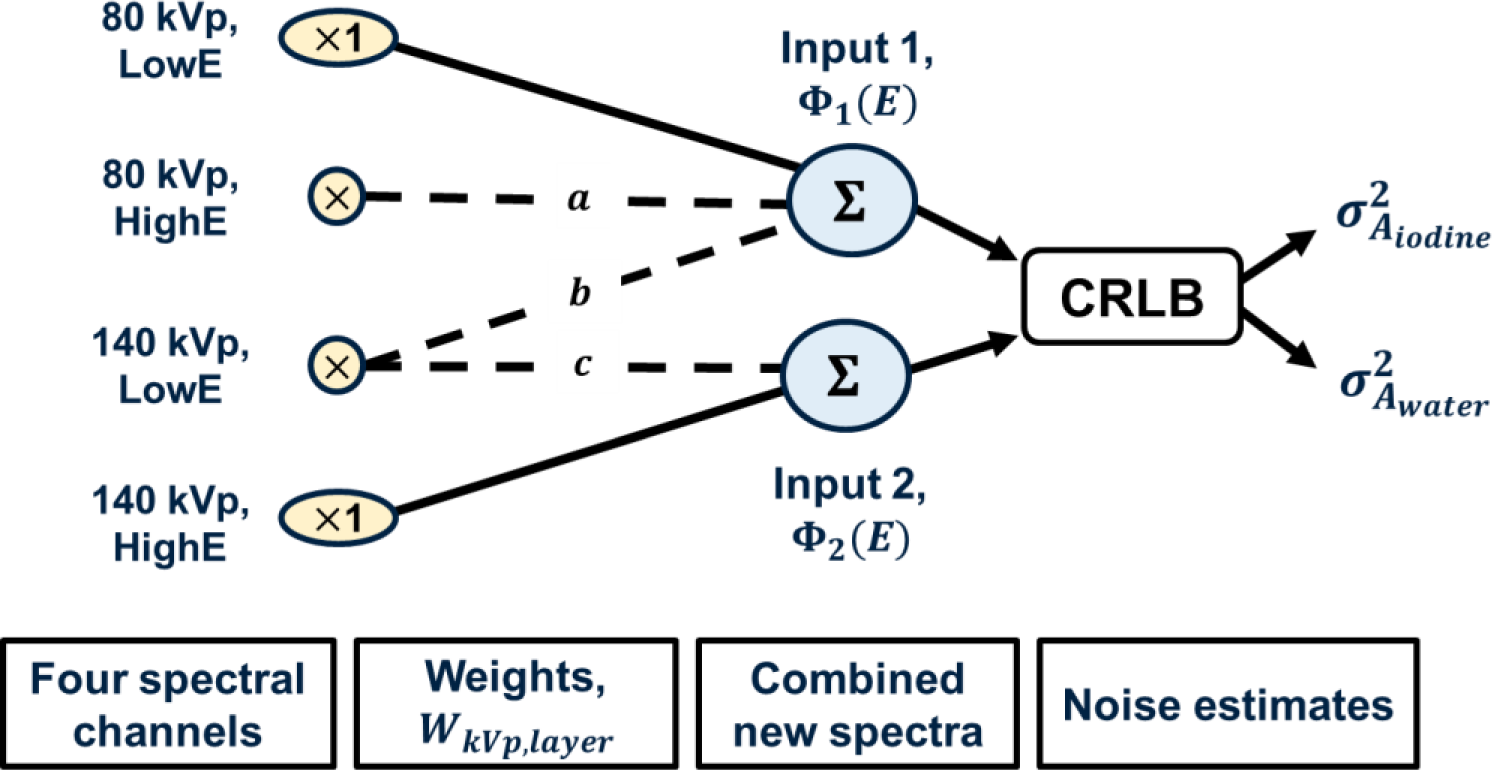
Outline of the methodology to reduce complexity of input dimensions through the linear combination of four channel inputs into two new spectra. The CRLB uses these two inputs (Φ_1_(*E*), Φ_2_(*E*)) based on weighting scheme and the weights (*a*, *b*, *c*) to estimate the noise in the projections for two basis materials, iodine and water.

We generated one final, additional dimensionally reduced weighting strategy by taking the minimal number of channels (2) using only the maximally spectrally separated data channels (*a, b, c = 0*) or W_1_ = [1,0,0,0] and W_2_ = [0,0,0,1]. While dose efficiency of this combination is low, it is instructive to explore the impact of spectral separation versus photon fluence on material decomposition performance using this method compared to kVp-s and weighting schemes 1 and 2.

### Performance statistics

In this paper, we utilize the Cramer-Rao lower bound of variance to estimate the noise on an iodine/water decomposition task through a simple phantom. Spectral CT reconstructions attempt to recover the line integrals, A_k_, for each projection, where k represents iodine or water.

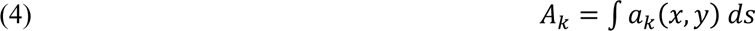

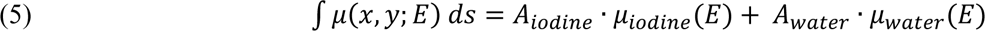

We derived the CRLB to compare a four-channel estimator performance to the simplified weighted two-channel estimator. In the four-channel estimation of noise, the four spectra *S*_*kVp*,*layer*_(*E*) are assumed to be independent signals. In the dimensionally reduced approach, new weighted spectrums Φ_1_(*E*) and Φ_2_(*E*) are considered independent signals. For simplicity and to generalize both estimators, we refer to all these spectra as *S*(*E*) in the following equations. We assumed Gaussian noise on each measurement, *m*_*j*_, representing the *j*^*th*^ total energy deposited in *j* = 1, …, *N* measurements with mean (*θ*) and variance (*σ*^2^). In the two-channel scenarios, we scaled the variance of the input signals Φ_*i*_(*E*) with respect to the square of each element in 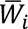 (7). This was necessary given the noise modeled must reflect the relative contribution of each data channel to the final signal.

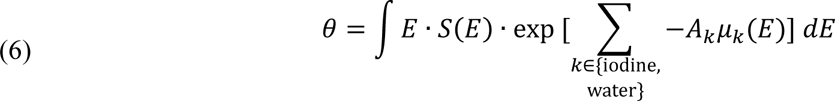

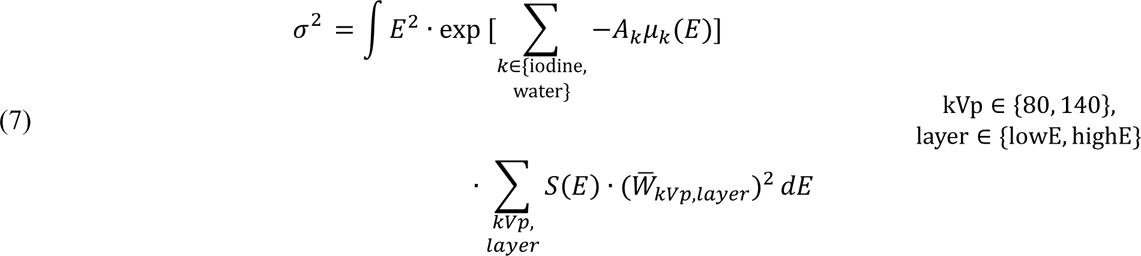

The likelihood of obtaining the actual measurement, *m*_*j*_, can be written as such:

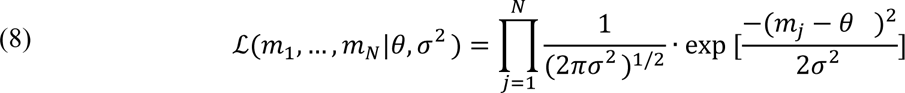

The negative log likelihood follows:

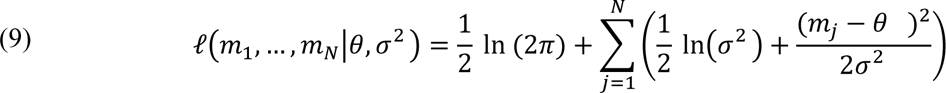

Because the negative log likelihood is dependent on *A*_*iodine*_ and *A*_*water*_, we can compute the Fisher information matrix, *I*(*θ*), by taking the expectation of the partial derivative of all likelihood functions with respect to *A*_*iodine*_ and *A*_*water*_.

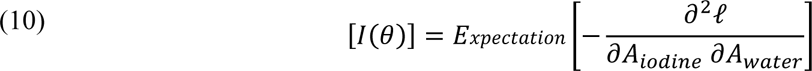

The Cramer-Rao lower bound states for an unbiased estimator that the lower bound of variance, assuming it to be finite is:

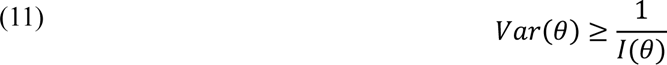

In the pure kVp-switching scenario, the input weight matrices 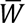 were [1, 1, 0, 0] and [0, 0, 1, 1] to represent the full utilization of the available 80 kVp layer information as one input to the decomposition and the full use of the 140 kVp layers as the second input. The visualization of this approach is shown in **Figure 3** on the left with the four-channel CRLB estimator approach schematic on the right. Although undesired to implement as a full material decomposition method with real data due to computational complexity and possible additional burden on the slip-ring, the four-channel estimator provided a baseline performance metric to compare different weighting strategies against. While both kVp-switching and the four-channel estimators use the full spectrum available, the difference is whether to combine layer signals before calculating the spectral noise. We computed the CRLB estimate of iodine and water noise for four-channel and the kVp-switching for each duty cycle and phantom size combination.

**Figure 3:**
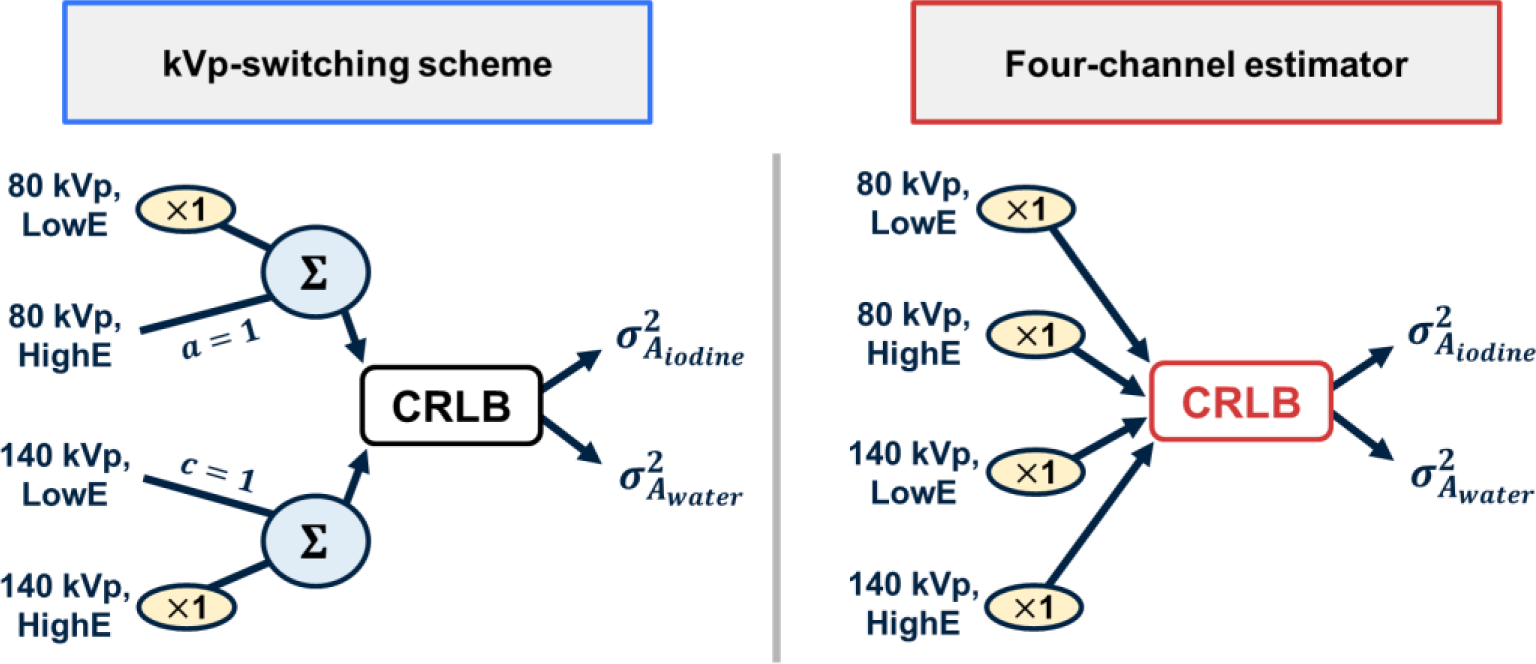
Graphical representation of the reduced dimension two-channel kVp-switching estimator weight scheme (left) and the four-channel CRLB (right) used to compute the CRLB noise estimates. Both approaches use 100% of the available spectral channels, but the kVp-switching scheme combines layer information prior to noise estimation.

### Weight optimization

To identify the values for *a, b* (scheme 1) and *a, c* (scheme 2) to minimize the CRLB noise estimates, we used the Python Scipy minimization function where the objective function was the iodine standard deviation noise estimate. The truncated Newton Conjugte-Gradient algorithm was used to iteratively update weight parameters to arrive at the minimum CRLB noise for all duty cycles and phantom sizes. An initialization weight of 0.2 was used for each variable. Valid weight values ranged from [0, 1].

## RESULTS

### Effect of photon energy ratios on duty cycle and estimated noise

We took a collection of 12 simulations using 300 mm of water and 25 mm of iodine with varying duty cycles from 0.1/99.9 to 90/10, including the five selected duty cycles (10/90, 25/75, 50/50, 75/25, 90/10), and normalized the tube spectra to contain the same total photon energy as a 50/50 duty cycle acquisition. **Figure 4** illustrates the relationship between fraction of high kVp photon energy (d_140_) relative to the total photon energy (d_140+80_) with the corresponding duty cycles and estimated relative CRLB iodine noise using kVp-switching (*a=1, b=0, c=1*) and the four-channel estimator. The CRLB noise has been normalized to the maximum noise produced by the kVp-s acquisition. The 10/90 duty cycle was closest to containing equivalent spectra photon energy between 140 and 80 kVp projections with the selected scan parameters. This dose ratio was where both kVp-switching and four-channel CRLB estimates were at their minimum across the selected ratios. Extreme photon energy ratios in either direction resulted in high estimated noise as the distribution of SNR between the two voltages’ spectra became highly unbalanced. Comparing kVp-s and four-channel noise estimates at different photon energy ratios, we noted that four-channel noise was always lower and more advantageous to use, especially at extreme duty cycle ratios. For example, using the 90/10 duty cycle, the four-channel iodine noise was nearly one-third of the predicted kVp-switching noise. This demonstrates the additional spectral separation capabilities of the four-channel detection. The comparison highlights the potential benefit a dual-layer detector may add to a kVp-s system.

**Figure 4:**
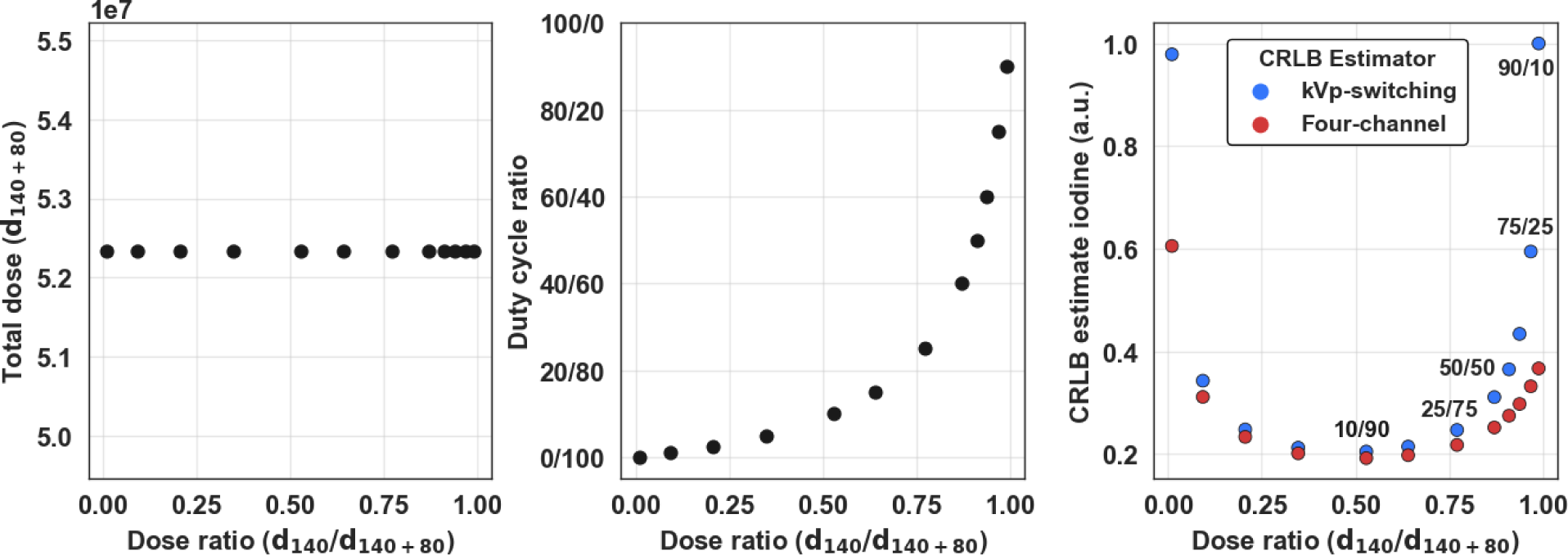
Demonstration of the effect of the ratio d_140_/d_140+80_ while d_140+80_ is held constant (left). The duty cycle experiences an exponential positive change as d_140_/d_140+80_ increases (center) while ratios near zero or one demonstrate high estimated noise for both kVp-s and four-channel estimator (right). A 50/50 ratio of d_140_/d_140+80_ roughly corresponds to a 10/90 duty cycle.

### Effect of duty cycle and weight values on CRLB noise estimate

Using 40 weights linearly spaced from 0 to 1 for each input weight, the CRLB estimation of iodine noise in the same 300 mm water phantom was plotted to examine the relationship between duty cycle, weighting scheme 1, and scheme 2. The noise values were normalized to the maximum CRLB output between both schemes using the 50/50 duty cycle so that a uniform scale could be visualized across the 10/90, 25/75, and 50/50 plots (**Figure 5**). The optimal weights for the lowest noise were then marked with a yellow star. **Figure 5** shows the clear impact of the duty cycle: as the 140 kVp component increases, the average normalized noise value in each plot increased going from 0.590 and 0.504, to 0.667 and 0.598, to 0.808 and 0.876 for schemes 1 and 2 respectively. Scheme 2 had the lower minimum CRLB iodine noise in the 10/90 and 25/75 duty cycles whereas scheme 1 generated the lowest expected iodine noise in the 50/50, 75/25, and 90/10 duty cycles. Across all duty cycles, the optimal weight for *a* ranged from 0.429 to 0.675 while all optimal *b* and *c* weights remained less than or equal to 0.153 and 0.327 respectively. In duty cycles 25/75 and increasing, the *c* weight to optimize the CRLB iodine noise was 0. In both schemes, as the 140 kVp component of the duty cycle increased, the optimal *a* value also increased.

**Figure 5:**
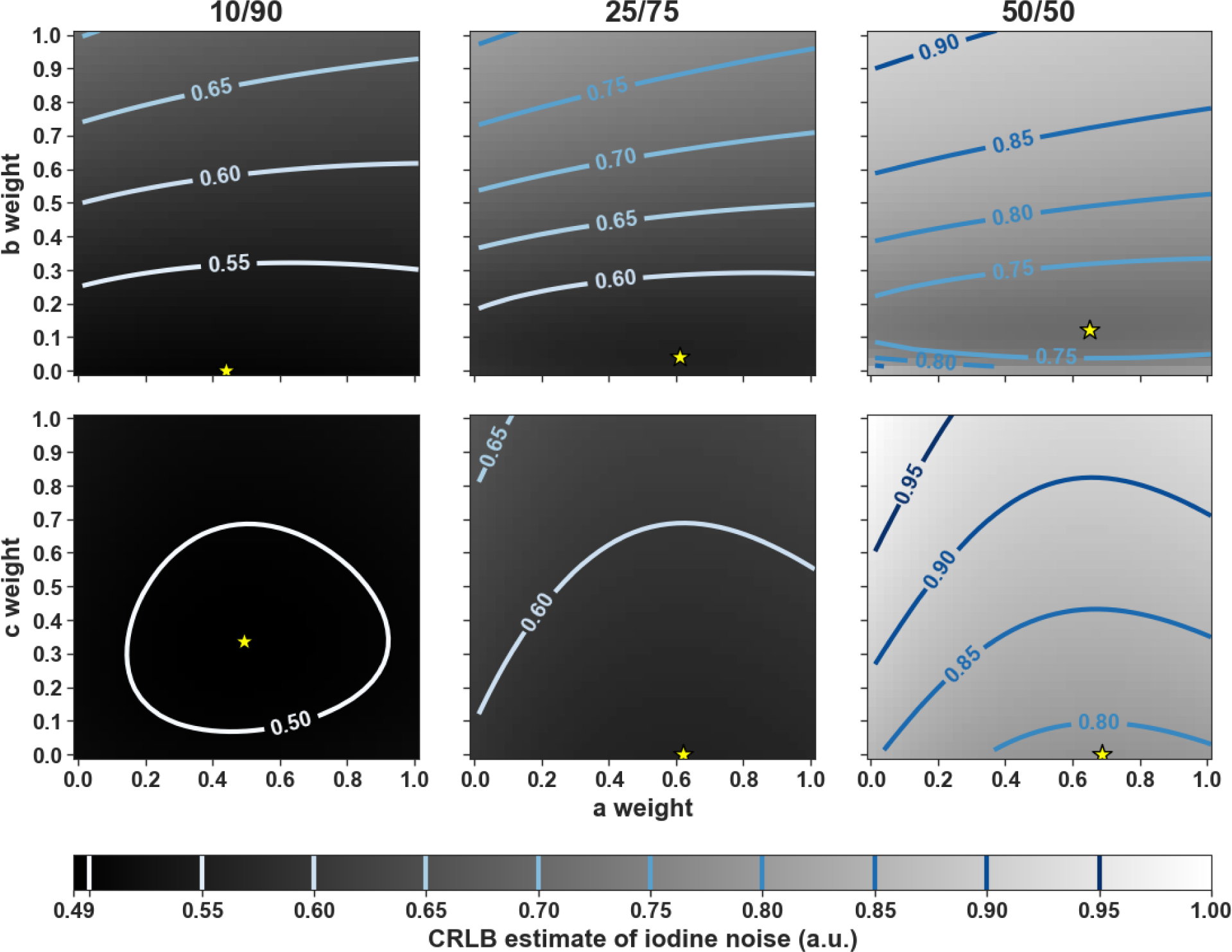
Using the 300 mm adult water length, we show the relationship comparing scheme 1 (upper row) and scheme 2 (lower row) weight values on estimated CRLB iodine noise relative to the max noise estimate in the 50/50 dataset. The yellow star indicates the weights with the lowest iodine noise estimate. In the 10/90 and 25/75 duty cycles, scheme 2 outperformed scheme 1, however scheme 1 had the lower iodine noise estimate in the 50/50, 75/25, and 90/10 duty cycles. Average noise estimates increased as the duty cycle included increased 140 kVp cycle time.

### Comparison of two-channel weighting schemes to the four-channel case with respect to noise estimate

The estimated iodine noise using the reduced dimension two-channel estimator schemes compared to the four-channel estimator were visualized for the 300 mm patient diameter (**Figure 6**). The bar in red represented the four-channel, ideal material decomposition noise performance given the tube spectra, detector response, and spectral separation at each duty cycle. This scheme contained the lowest noise across all other tested schemes for all duty cycles and patient thicknesses. As the 140 kVp component increased, the estimated noise for all schemes increased, although the magnitude varied for each scheme. From the 10/90 duty cycle to the 90/10 duty cycle, the four-channel noise nearly doubled while the kVp-s noise more than tripled. Within the 90/10 duty cycle simulations, the optimal weighting for scheme 2, kVp-switching, and the maximally separated weighted noise estimates were over 2x the estimated four-channel noise. However, the optimal weighting scheme 1 noise was 1.0027x greater than the four-channel estimate. The maximally separated reconstruction scheme CRLB estimate was always less than the kVp-switching noise and greater than both the optimal weighting schemes 1 and 2. This figure importantly demonstrates the ability for one of the two-channel weighting schemes, using optimal weights, to produce an estimated iodine noise within 0.27% of the four-channel noise. Scheme 2 again shows better performance than scheme 1 only in the 10/90 duty cycle.

**Figure 6:**
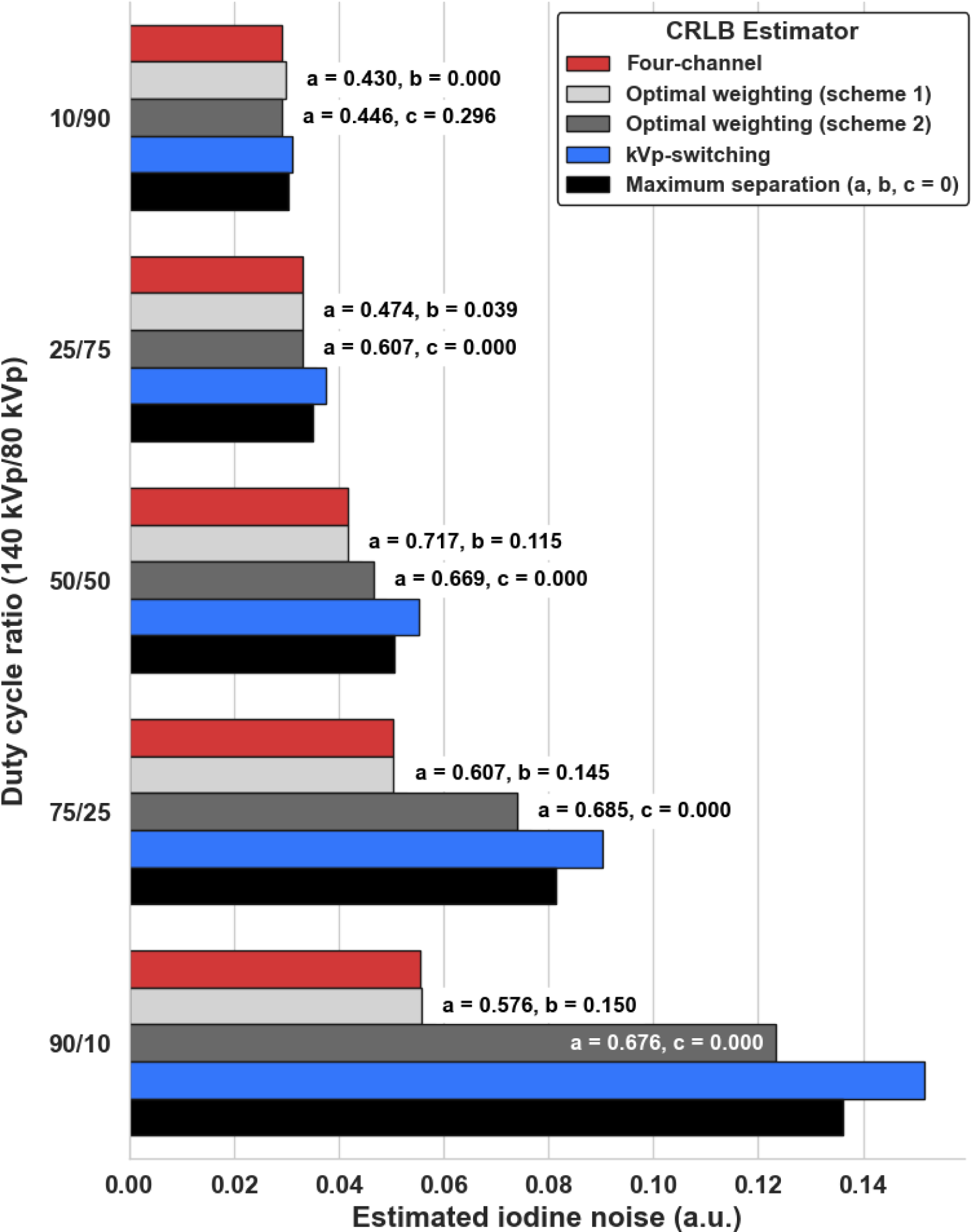
Comparison of two-channel CRLB iodine noise weighted schemes relative to the four-channel estimator noise for each duty cycle for the 300 mm patient size. As the 140 kVp component of the duty cycle increases, the noise estimates for kVp-switching noise and maximally separated weighting become larger. In comparison, the optimal weighting scheme, particularly scheme 1, remains robust and capable of achieving noise levels within 0.26% of the four-channel estimate, given by proximity to the red bar, regardless of duty cycle.

### Pediatric- and large adult-sized phantom weighting

For the 400 mm, large adult case, we found the best two-channel performance was the optimal weighting scheme 2 in the 10/90 duty cycle where the noise was only 0.05% greater than the corresponding four-channel noise. In the 90/10 duty cycle scenario, the best two-channel noise performance was scheme 1 with 1.018x the noise of the four-channel iodine noise. On average, kVp-s CRLB noise values were 1.536x greater than the ideal noise while scheme 1 noise was only 1.01x greater than the lowest noise estimates. Trends for the 150 mm, pediatric-sized phantom case were the same as observed for the other two phantom sizes. On average, optimal weighted scheme 1 iodine noise was 1.006x greater than the four-channel estimates for the same duty cycles. In comparing these different water path lengths, we demonstrated that the weighting system can receive specific patient parameters and generate optimal weights to produce noise estimates similar in magnitude with the ideal four-channel material decomposition estimator.

## DISCUSSION

We introduced a computationally efficient method to address data dimension reduction necessary in multi-channel spectral CT systems. Our ultimate goal for such a system is to maximally reduce bias in material decomposition, particularly of iodine contrast because accurate iodine quantification and visualization increases the screening and diagnostic utility of CT imaging. These simulation studies can be easily modified to fit more diverse body sizes, iodine concentrations, and scan parameters including tube voltage, tube current, and duty cycle. Most importantly, they provide a feasible solution to optimally use available data and to integrate with fast and efficient material decomposition methods. We show that our weighting system estimates an improved iodine noise performance compared to kVp-switching alone. Finally, we show that the performance of these weighted reduced dimension two-channel systems matches the performance of a four-channel estimator.

The optimal weights determined by the minimization of CRLB iodine estimated noise reveal how to best utilize the additional spectra data by balancing photon fluence and the desirable photon energy separation. Given scheme 1 produces the lowest noise in all tested duty cycles but 10/90 and 25/75, we show that high kVp, low energy photon information from the upper detector layer combined with the low kVp projections creates a more ideal spectral input rather than combining the layer data with the 140 kVp when switching parameters favor the 140 kVp tube voltage component. We replicated these findings for multiple simulated patient sizes.

The use of the CRLB to predict performance of material decomposition from spectral inputs has been described by Roessl and Hermann^30^ and Yang et al.^31^ using both dual-layer and photon-counting detector technology. Though we do not model every system imperfection (crosstalk, K-edge escape, scatter, etc.), we used a precise physical model that captures primary spectral effects which directly play a role in material decomposition. Future research with available CT instrumentations will require application of weights to fully integrate non-linear, imaging chain effects not captured in this simplified simulation. We will also explore optimization of other quantification tasks including monoenergetic signal-to-noise and contrast-to-noise performance.

While we selected one iodine concentration and only five duty cycles for testing hybrid kVp-s and dual-layer acquisitions, we recognize that additional clinical input may be required to define realistic protocols. The cycle time intervals are heavily dependent on both tube and detector integration period limitations which vary for different vendor components. For some specific scanners, there have been findings on setting ideal flux ratios between 80 and 140 kVp^35^. From a tube and generator perspective, it is desirable to maintain a high tube current with a short 140 kVp duration to create a short transition time from 140 kVp to 80 kVp because this transition is mainly driven by the built-in tube current discharging and parasitic capacitors. A faster transition time would help eliminate intermediate spectra occurring between the high and low tube voltages and improve spectral separation. At the detector, reducing 140 kVp cycle time relative to 80 kVp duration is important as sufficient flux from the 80 kVp spectra is necessary to avoid photon starvation and to reduce the impact of electronic noise. From a patient perspective, minimization of 140 kVp cycle time is also important to reduce extended radiation dose exposure to high energy photons. We define a simple case for this simulation study but will seek to address these clinical and equipment concerns in future experimental implementation of this method.

Finally, we note that this process could be replicated for a CT with a photon-counting detector and potentially other tube configurations. Yang et al. demonstrated an eight-bin compression strategy for a silicon-based photon-counting detector that again capitalized on the CRLB to find ideal weighting combinations^36^. We chose to show our weighting method by using a rapid kVp-switching x-ray tube and dual-layer detector hybrid system. This configuration leverages the inherent high spectral separation of incident spectra while further discriminating the high kV and low kV acquisitions into low energy and high energy photon domains produced from the dual-layer detector. Experimental work acquired on clinical analog systems will be required to validate this method for managing and optimizing multi-channel spectral data.

## CONCLUSION

In this paper, we demonstrated a strategy for optimal dimensionality reduction of spectral data acquired on multi-channel spectral CT systems by leveraging an analytic statistical tool. By combining spectral technologies, we doubled the number of spectral channels, creating four possible distinct photon energy inputs to use in material decomposition. We compared a statistical estimation of noise in the iodine domain between the maximum likelihood estimator of variance using all four channels compared to a simplified two-input CRLB estimator. We showed that estimated material decomposition noise performance from a simplified weighting scheme using optimal weights could achieve noise as low as a theoretical four-dimension material decomposition. We also illustrated that this noise is lower than the noise estimated in a kVp-switching protocol with the same spectra regardless of duty cycle. This provides evidence that multi-channel spectral CT improves spectral performance for two material decompositions, e.g., addition of a dual-layer detector improves spectral performance of a kVp-switching system specifically for iodine estimation. It highlights how inclusion of more spectrally distinct and diverse data may improve material decomposition.

In CT diagnostic imaging, the use of iodine contrast is paramount for visualization of vascular processes and pathologies. This task becomes exceedingly difficult at lower radiation doses or with use of smaller iodine volumes and concentrations, both which are generally always desired for increased patient safety^37^. Thus, there is demand for a spectral CT system which can accommodate low concentration iodine, low radiation dose exposure, and delivery of accurate iodine quantification. Combining existing technologies or using state-of-the-art hardware has been theorized to fulfill these requirements, but optimization of these systems to utilize increased spectral information, in a clinical feasible timeframe, has yet to be fully explored. We demonstrate a flexible method to incorporate multi-channel spectral information in an optimized weighting configuration to produce ideal iodine quantification results across a diverse range of patient sizes.

## Data Availability

All data produced in the present study are available upon reasonable request to the authors

## ACKNOLWEDGEMENT

We acknowledge support through the National Institutes of Health (R01EB030494).

